# HIV co-infection is associated with lower tuberculosis bacterial burden independent of time to diagnosis in Botswana, a setting with widespread ART use

**DOI:** 10.1101/2022.03.28.22273071

**Authors:** Juliana S. Chalfin, Chelsea R. Baker, Balladiah Kizito, Dimpho Otukile, Matsiri T. Ogopotse, Sanghyuk S. Shin, Chawangwa Modongo

**Author notes:** Corresponding author: Sanghyuk Shin, PhD, Associate Professor, Sue & Bill Gross School of Nursing, Director, UCI Infectious Disease Science Initiative, University of California Irvine, 106F Berk Hall Irvine, CA 92697. Co-first author (Juliana S. Chalfin, Chelsea R. Baker). Co-senior author (Sanghyuk S. Shin, Chawangwa Modongo).

## Abstract

HIV co-infection has been shown to be associated with lower tuberculosis (TB) bacterial load in studies conducted prior to widespread availability of antiretroviral therapy (ART). We investigated associations between HIV co-infection and TB bacterial load, accounting for differences in time to TB diagnosis, in a high prevalence setting with widespread ART use. In Gaborone, Botswana, 268 sputum samples from people with newly diagnosed TB were tested with Xpert MTB/RIF Ultra (Xpert). TB bacterial load and time to TB diagnosis were estimated using mean Xpert cycle threshold (CT) and symptom duration, respectively. Multiple linear regression models and causal mediation analysis were used to determine the associations between HIV and Xpert CT and assess the mediating effect of symptom duration. Mean CT values were higher in people living with HIV compared to people without HIV (22.7 vs 20.3, p < 0.001). Among those living with HIV, there was a negative relationship between CD4 count and mean CT value (Spearman’s rho -0.20, p = 0.06). After controlling for gender, age, and symptom duration, HIV status remained associated with CT value, with an average increase of 1.6 cycles (p = 0.009) among people with HIV and CD4 count > 200 cells/mm^3^ and 2.1 cycles (p = 0.002) in those with a CD4 count ≤ 200 cells/mm^3^ compared to individuals without HIV. Symptom duration was also found to be associated with CT value (p < 0.05). We found an indirect effect of HIV status on Xpert CT through the mediator, symptom duration (β = 0.33, p = 0.048), accounting for 13.5% of the relationship. Our findings suggest that time to TB diagnosis partially mediates the relationship between HIV status and CT value, but differences in pathophysiology between people with and without HIV likely play a dominant role in affecting TB bacterial burden.

## Introduction

Globally, tuberculosis (TB) has remained a leading cause of death due to an infectious disease with an estimated incidence of 10.0 million people with TB and 208,000 TB deaths among people living with HIV (1). While the World Health Organization (WHO) has provided goals to end the global TB epidemic, recent data indicate that targets for reduced incidence rates and deaths are not being met (2). In high burden areas, HIV poses as the greatest risk factor for developing TB, exacerbating existing susceptibility to primary infection or reactivation of latent TB (3). Southern Africa continues to report the highest prevalence of TB/HIV co-infection, demonstrating an urgent need to better understand TB transmission dynamics to reduce TB incidence and mortality (4).

The likelihood of TB transmission depends in part on the degree of infectiousness, which is often measured in clinical settings using bacterial load present in sputum (5). While this is commonly done using smear microscopy, Xpert MTB/RIF assays (Xpert) have been suggested as an alternative (6,7). Xpert is a diagnostic test that uses real-time polymerase chain reaction (PCR). Quantitative estimation of TB bacterial load can be inferred through cycle threshold (CT) values (8). CT values convey an inverse relationship to bacterial load as a higher CT value signifies lower bacterial load and lower CT value signifies higher bacterial load.

Sputum bacterial load is influenced by pathophysiology of active TB disease, which involves destruction of lung tissue, generating cavities that pass *M. tuberculosis* bacteria to airways (9). Therefore, the level of lung damage contributes to the transmission potential of a TB-infected individual. However, among people with HIV and TB, lower CD4+ T cell counts have been associated with reduced lung inflammation (9). The reduced inflammation can result in decreased cavitation, sputum bacterial load, and ultimately reduced infectiousness, as suggested by the higher frequency of smear-negative pulmonary TB within untreated people living with HIV (9,10). For example, a previous study has corroborated the findings that increased immunosuppression, defined as CD4 count < 200 cells/mm^3^, was associated with decreased bacterial burden, measured through CT values (6). Consequently, untreated HIV infection can change TB disease presentation and reduce TB infectiousness.

Bacterial burden may be impacted by delays in diagnosis, as the active disease state involves bacilli replication. Decreased TB bacterial burden with HIV co-infection may be partially related to early TB diagnosis and treatment initiation (9,10). Treatment for HIV typically involves frequent contact with the healthcare system, including increased opportunities for TB case finding and prompt treatment (9). Therefore, the association between HIV status and TB bacterial burden may be mediated by earlier TB diagnosis among people living with HIV.

Antiretroviral therapy (ART), the treatment of choice for managing HIV/AIDS, has significantly improved the quality of life of people living with HIV since it decreases the viral load and restores normal immune function against opportunistic pathogens (11). Access to effective treatment for HIV has greatly expanded, and as of 2020 an estimated 75% of people living with HIV (with known HIV status) were receiving ART (12). Further research is needed to understand its effect in settings with a high burden of both HIV and tuberculosis (9). As ART alters the disease process of HIV, there is a gap in knowledge of whether HIV-coinfection remains associated with reduced TB infectiousness during the present time when ART is widely available.

The purpose of this study was to determine associations between HIV co-infection and sputum TB bacterial load and assess whether differences in duration of symptoms mediates this association, in a high prevalence setting. This study could contribute to understanding how HIV-coinfection affects infectiousness and transmission risk of TB in the current era of widespread ART use.

## Methods

### Study Design, Setting, and Population

Botswana is a country in southern Africa with a high burden of TB and among the highest rates of TB-HIV co-infection in the world. This cross-sectional analysis is part of an ongoing population-based TB transmission study in Gaborone, the capital city of Botswana. Participants were sequentially enrolled after diagnosis of TB from community or health care facilities. All participants were recruited in accordance with Botswana national guidelines. Active case finding, involving the screening of all patients, and passive case finding, involving clinical diagnosis of patients presenting with TB symptoms, were utilized in 26 public health facilities. Participants included male and female patients of all ages. Clinical and demographic data were collected through a standardized questionnaire that captured information about TB symptoms and their duration (< 1 month, 1-2 months, 2-3 months, and > 3 months prior to diagnosis). Smoking and alcohol use were collected as these variables are known to impact TB transmission as well as the immune system. Smoking has been shown to increase infectiousness by increasing the aerosolization of bacilli through changes in mucus and increased cough frequency (13). Furthermore, alcohol impairs the immune system leading to increased susceptibility to TB as well as reactivation and is associated with social environments that facilitate TB transmission (14). Participants provided their consent for us to access their medical records to collect TB and HIV history, including CD4 cell counts, viral load, and ART history. We recorded CT values for five probes in the Xpert (Xpert MTB/RIF Ultra) test used for clinical diagnosis. Mean CT values across all probes was used as the primary measure of bacterial load. Participant-reported duration of symptoms was used as an approximate measure of time to diagnosis.

### Statistical Analysis

Standard descriptive statistics were used to characterize the data. The sample population of CT values was compared between groups of participants by HIV status and CD4 count using the nonparametric Wilcoxon test. CT values were also compared between groups of participants by symptom duration using the Wilcoxon test to determine significant difference. Odds ratios were estimated using a dichotomous variable for symptom duration (< 1 month and ≥ 1 month) by HIV status and CD4 count category. Spearman correlation was used to assess the relationship between CD4 counts and mean CT values among participants living with HIV since values were not normally distributed. Bivariate linear regression was used to examine the association between mean CT value as the outcome variable and selected patient characteristics as predictor variables. Multivariate linear regression models were used to control for confounding variables including age and gender to examine the relationship with HIV status as the primary exposure variable of interest. We also compared multivariate models with and without symptom duration to estimate the extent to which the association between HIV status and bacterial load is explained by differences in time to diagnosis. Causal mediation analysis was performed using the R package mediation to determine the mediation effects of symptom duration, as a dichotomous variable, on the relationship between HIV status and CT value (15). Bootstrapping was used to test the significance of the indirect effect, using 500 samples. All statistical analyses were conducted using the R statistical software version 4.1.2 (16).

### Ethics Approval

This study was approved by institutional review boards at the University of Irvine, California and Botswana Ministry of Health and Wellness Human Research Development Committee. All participants provided written consent for research use of clinical data.

## Results

### Sample Population

During January 2021 to August 2022, 379 participants had positive Xpert results, of which 338 participants had valid corresponding Xpert CT value results. Of those, 70 individuals (20.7%) were excluded from analysis due to missing HIV results (n=12), CD4 counts that were either missing (n=42) or taken greater than six months from the initial interview (n=13), or missing symptom duration (n=3). A total of 268 participants were included in our analysis.

Overall, 68.7% were male, and the median age was 34 (interquartile range [IQR]: 25-45.25) (Table 1). People with HIV comprised 33.2% with a CD4 count median of 263 cells/mm^3^ (IQR: 100-523). At the time of enrollment, 25% and 43.3% reported smoking and alcohol use, respectively. The proportion of participants reporting TB symptoms for < 1 month was 55.1% and 38.5% among people with and without HIV, respectively. Among individuals with HIV, 43.8% had CD4 counts of ≤ 200 cells/mm^3^, 61.8% reported taking ART at the time of TB diagnosis, and median viral load was 30.0 copies/mL (IQR: ≤20-400; Figure 1).

**Table 1:** Demographic and Clinical Information by HIV Status of Individuals in Botswana 2021-2022.

|  | Negative<br>(N=179) | Positive<br>(N=89) | Overall<br>(N=268) |
| --- | --- | --- | --- |
| Gender |  |  |  |
| Female | 57 (31.8%) | 27 (30.3%) | 84 (31.3%) |
| Male | 122 (68.2%) | 62 (69.7%) | 184 (68.7%) |
| Age |  |  |  |
| 0-19 | 18 (10.1%) | 0 (0%) | 18 (6.7%) |
| 20-29 | 72 (40.2%) | 10 (11.2%) | 82 (30.6%) |
| 30-39 | 42 (23.5%) | 29 (32.6%) | 71 (26.5%) |
| 40-49 | 19 (10.6%) | 28 (31.5%) | 47 (17.5%) |
| 50-65 | 17 (9.5%) | 22 (24.7%) | 39 (14.6%) |
| 65+ | 11 (6.1%) | 0 (0%) | 11 (4.1%) |
| Currently Smoking |  |  |  |
| Yes | 44 (24.6%) | 23 (25.8%) | 67 (25.0%) |
| No | 135 (75.4%) | 66 (74.2%) | 201 (75.0%) |
| Currently Drinking Alcohol |  |  |  |
| Yes | 78 (43.6%) | 38 (42.7%) | 116 (43.3%) |
| No | 101 (56.4%) | 51 (57.3%) | 152 (56.7%) |
| Symptom Duration |  |  |  |
| <1 month | 69 (38.5%) | 49 (55.1%) | 118 (44.0%) |
| 1-2 months | 70 (39.1%) | 27 (30.3%) | 97 (36.2%) |
| 2-3 months | 16 (8.9%) | 9 (10.1%) | 25 (9.3%) |
| 3+ months | 24 (13.4%) | 4 (4.5%) | 28 (10.4%) |
| HIV Status |  |  |  |
| HIV- | 179 (100%) | 0 (0%) | 179 (66.8%) |
| HIV+ CD4 > 200 | 0 (0%) | 50 (56.2%) | 50 (18.7%) |
| HIV+ CD4 ≤ 200 | 0 (0%) | 39 (43.8%) | 39 (14.6%) |

**Figure 1:**
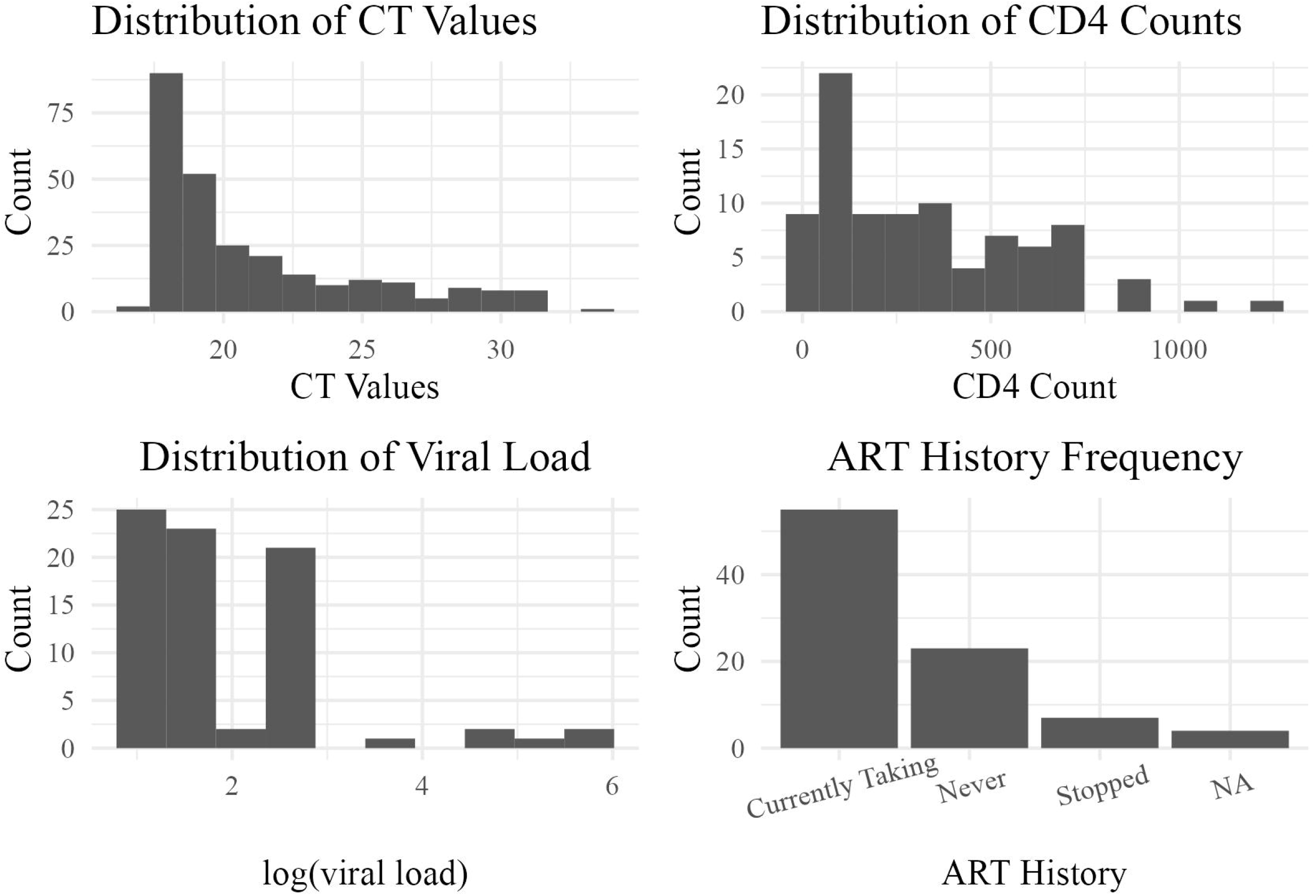
(A) Histogram of individuals’ mean CT values among all participants. (B) Histogram of individuals’ CD4 counts among participants living with HIV. (C) Histogram of individuals’ viral load among participants living with HIV. (D) Bar graph of frequency, measured in percent, of ART history among participants living with HIV.

### Symptom Duration

The percentage of participants experiencing symptoms >1 month was 61.5% among people without HIV, 48% among those with HIV and CD4 counts > 200 cells/mm^3^, and 41% among those with HIV and CD4 counts ≤ 200 cells/mm^3^ (Figure 2). Participants living with HIV with CD4 counts ≤ 200 cells/mm^3^ had significantly decreased odds of symptom duration >1 month compared to those without HIV (OR 0.44, p = 0.021), but no statistically significant difference was found between people with HIV and CD4 count >200 cells/m3 and people without HIV (Table 2).

**Table 2.**
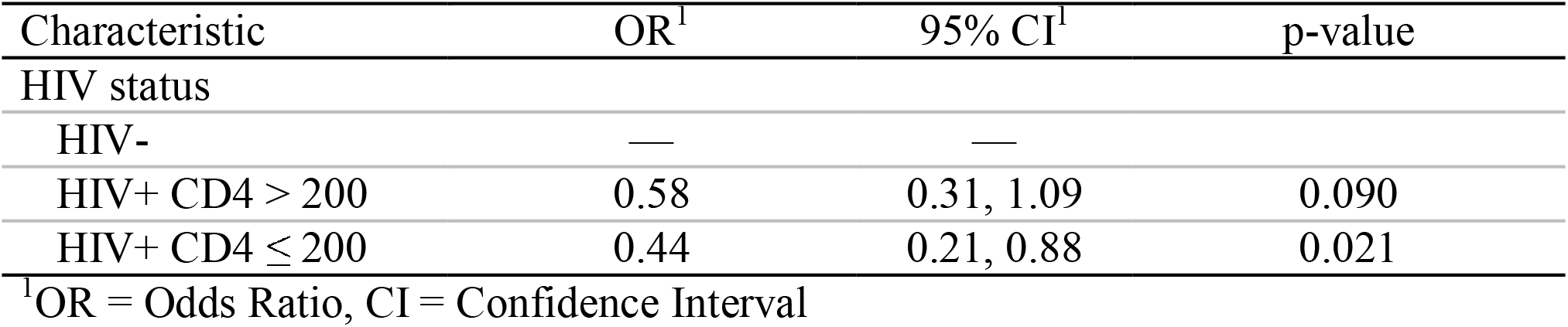
Odds ratio for Symptom Duration Greater than 1 Month by HIV Status and CD4 Count Breakdown.

**Figure 2:**
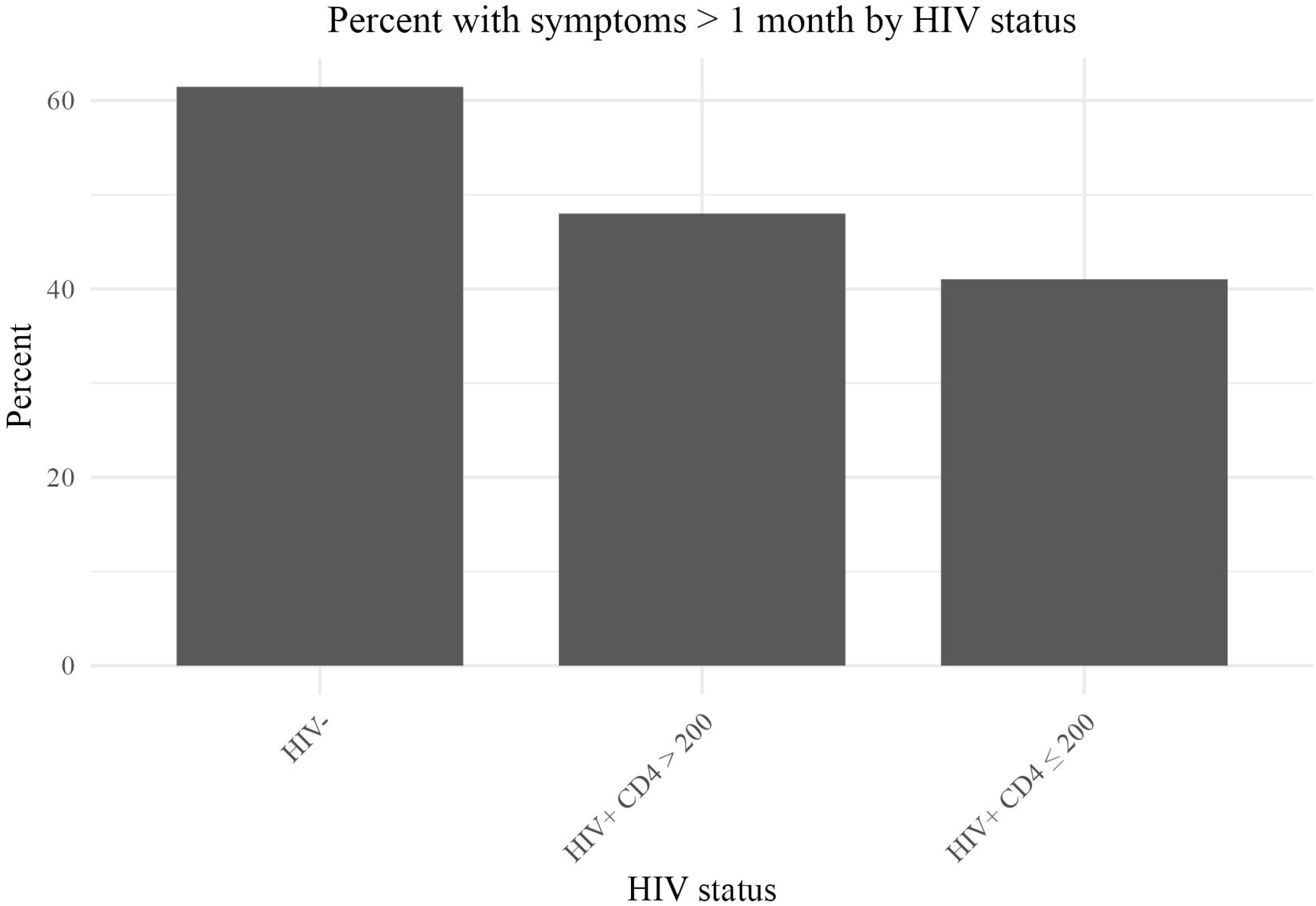
Comparison of symptom duration length between HIV status.

### Distribution of CT Value

The mean CT value across all participants was 21.1 (IQR: 18.1-23.1) with a range of 17.1-33.9 (Figure 1A). Mean CT values were higher among people living with HIV in comparison to people without HIV (22.7 vs 20.3, p < 0.001) (Figure 3A). This was noted for those with CD4 >200 cells/mm^3^ (p = 0.009) as well as ≤ 200 cells/mm^3^ (p < 0.001). However, the difference between participants in the two CD4 count categories was not significant (23.0 vs 23.3 for CD4 >200 cells/mm^3^ and ≤ 200 cells/mm^3^, respectively, p = 0.34).

**Figure 3:**
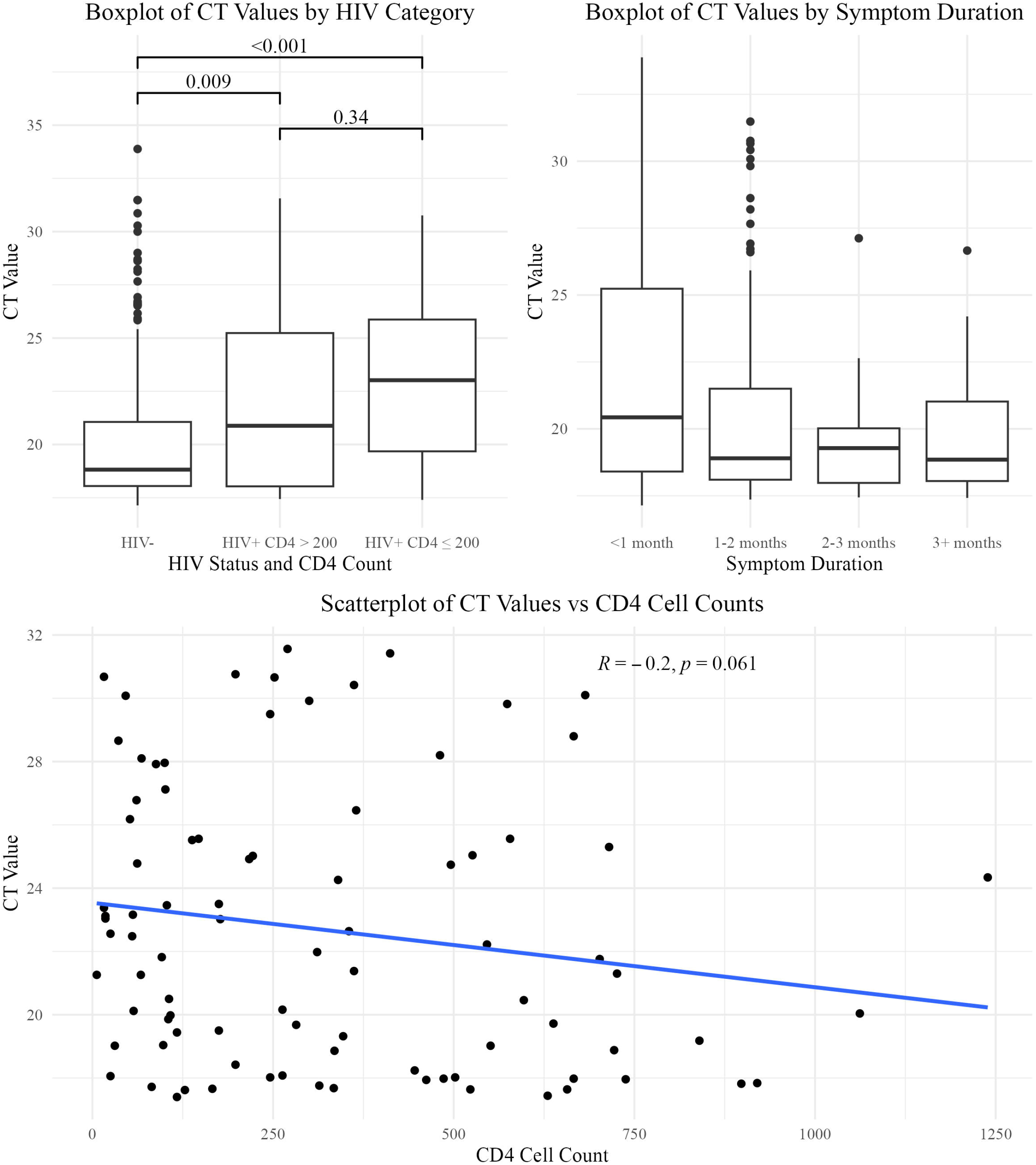
(A) Boxplot comparing mean CT values between HIV status and CD4 count, among those living with HIV. (B) Boxplot comparing mean CT values between groups of symptoms duration prior to diagnosis, measured in months. (C) Scatterplot comparing mean CT values and CD4 cell counts among participants with HIV.

Among participants living with HIV, there was a weak negative correlation between CD4 count and mean CT value (Spearman’s rho -0.2; p = 0.06; Figure 3C). Participants who reported symptoms for < 1 month had higher mean CT values compared to those that reported symptoms for 1-2 months, 2-3 months, and > 3 months prior to diagnosis (p = 0.013; p = 0.013; p = 0.017 respectively) (Figure 3B).

### Individual Level Predictors of CT Value

In a bivariate linear regression model, compared to participants without HIV, mean CT values were increased by 2.1 (p < 0.001) in participants living with HIV with CD4 counts >200 cells/mm^3^ and by an average of 2.7 (p < 0.001) in participants living with HIV with CD4 counts ≤ 200 cells/mm^3^ (Table 3). The bivariate linear regression model displays a decreasing trend in mean CT values as the duration of symptoms increases.

**Table 3.**
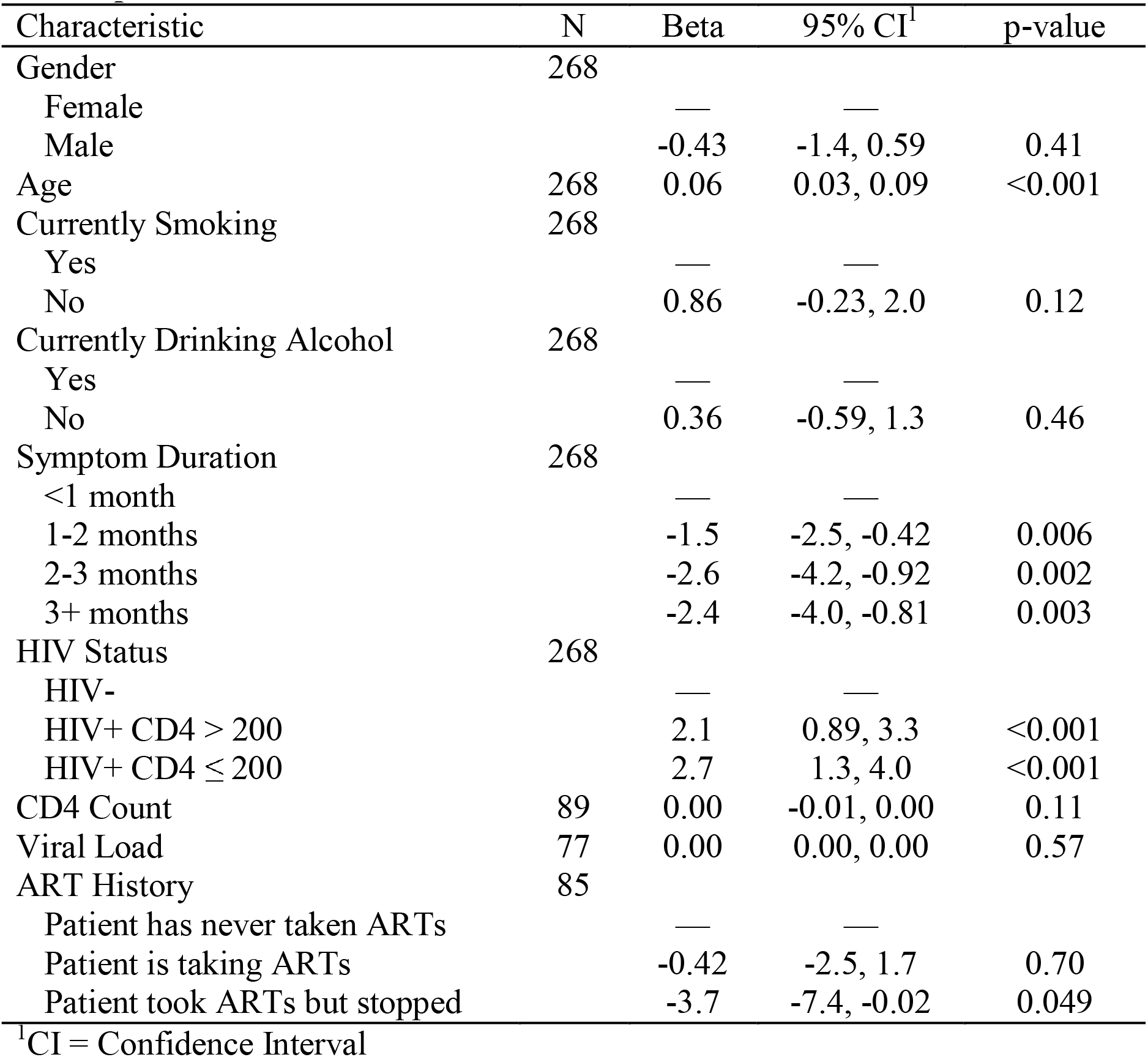
Bivariate Linear Regression Model for Mean CT Value among All Participants.

multivariate linear regression model, HIV status, including the CD4 count breakdown for In a participants with HIV, and symptom duration were significant predictors of mean CT (Table 4). When controlling for gender, age, and symptom duration, the mean CT value was 1.6 cycles higher (p = 0.009) among people with HIV and CD4 count > 200 cells/mm^3^ and 2.1 cycles higher (p = 0.002) in those with a CD4 count ≤ 200 cells/mm^3^ compared with the reference group of individuals without HIV. After controlling for gender, age, and HIV status, the mean CT value decreased by 1.1 cycles (p = 0.026), 2.2 cycles (p = 0.007), and 1.6 cycles (p = 0.043) for participants that reported TB symptoms 1-2 months, 2-3 months, and > 3 months, respectively, prior to diagnosis compared to those that reported symptoms < 1 month prior. The mean CT value also increased by 0.04 cycles with age as a continuous variable (p = 0.022) in the multivariate linear regression model controlling for gender, HIV status, and symptom duration. In a multivariate linear regression model that only included people with HIV, the association between CD4 count and CT did not reach statistical significance when controlling for gender, age, CD4 count, viral load, and ART History (p= 0.2; Table 5).

**Table 4.**
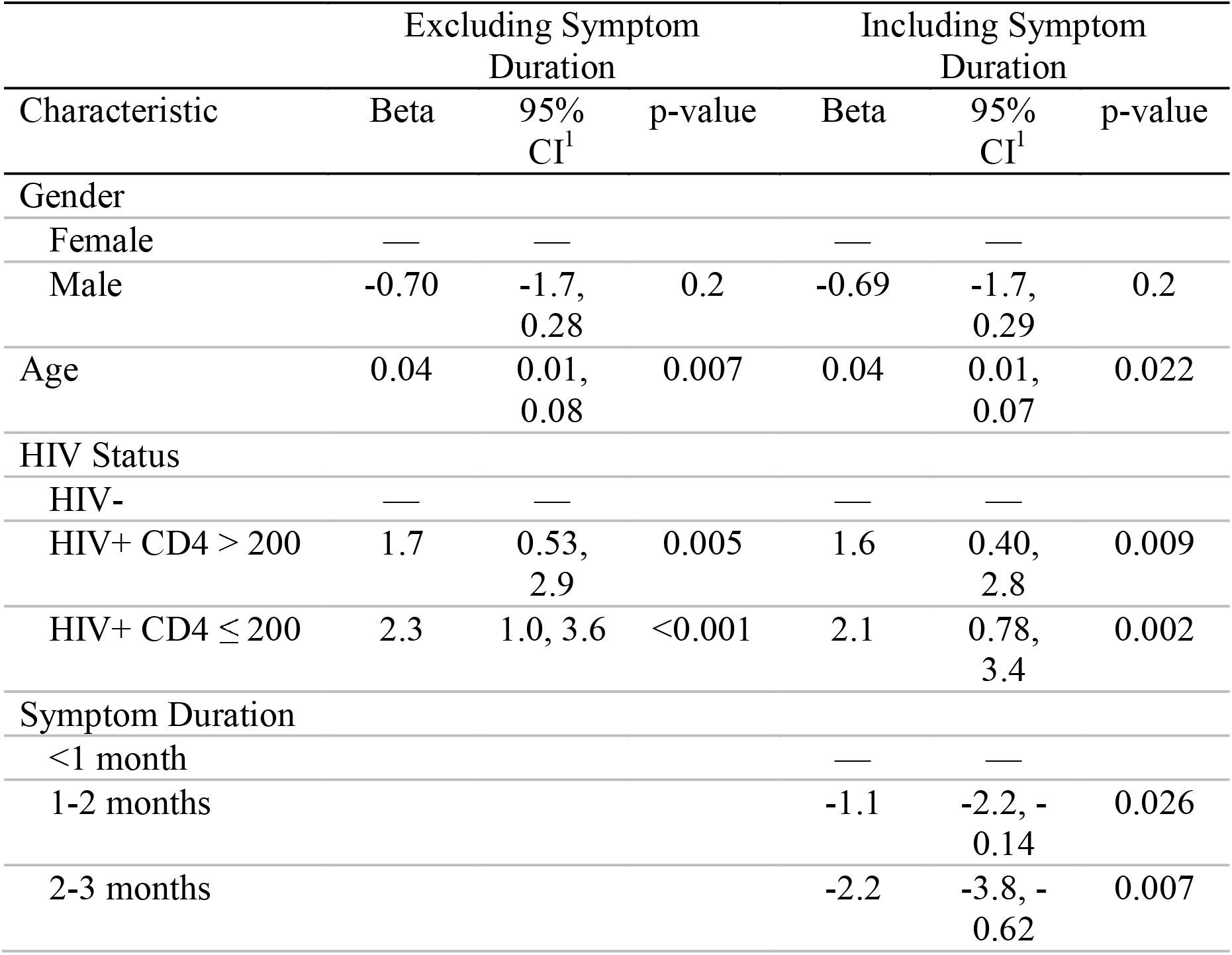
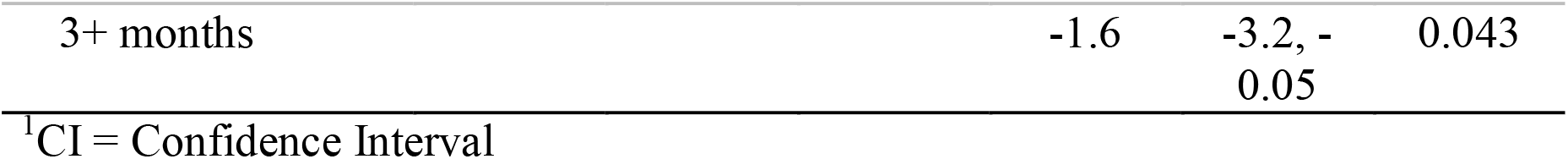
Multivariate Regression Models for Mean CT among All Participants.

**Table 5.**
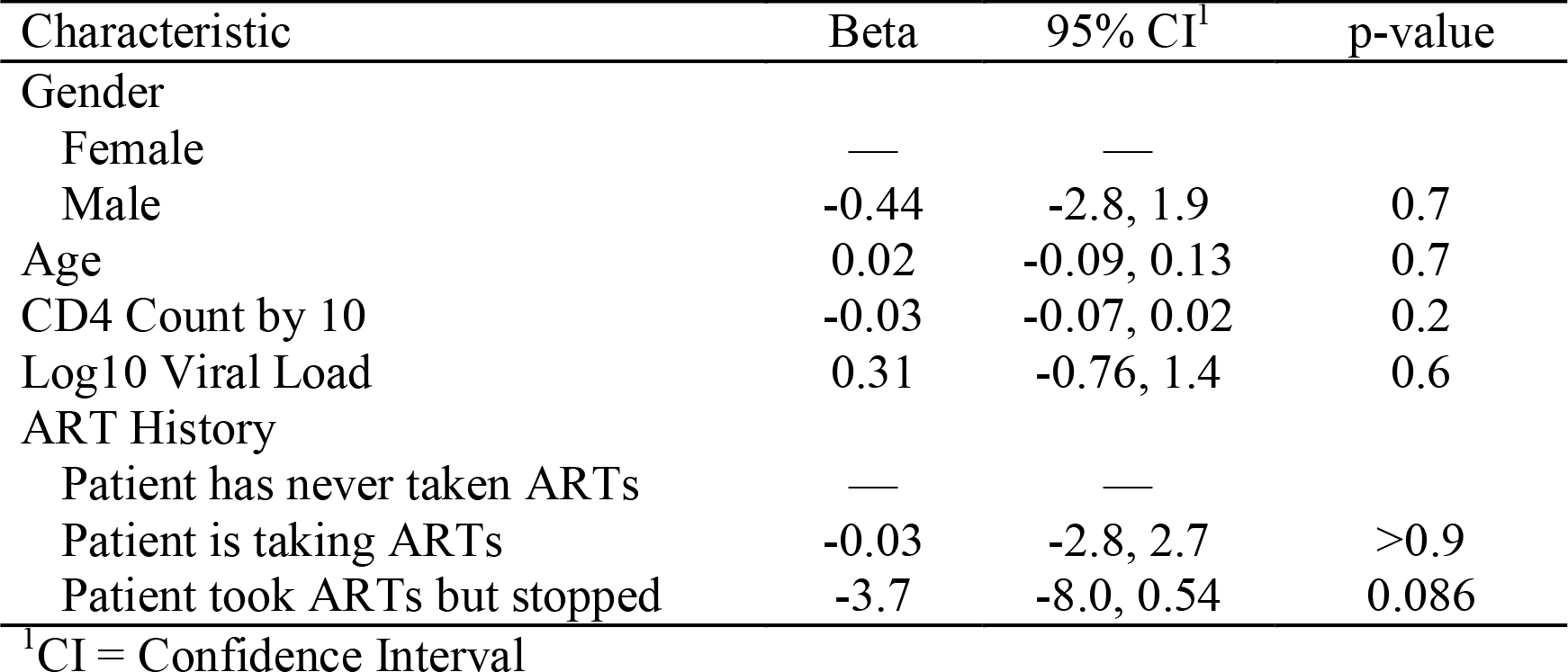
Multivariate Regression Model for Mean CT among Participants Living with HIV.

| Characteristic | Beta | 95% CI<sup>1</sup> | p-value |
| --- | --- | --- | --- |
| Gender |  |  |  |
| Female | — | — |  |
| Male | -0.44 | -2.8, 1.9 | 0.7 |
| Age | 0.02 | -0.09, 0.13 | 0.7 |
| CD4 Count by 10 | -0.03 | -0.07, 0.02 | 0.2 |
| Log10 Viral Load | 0.31 | -0.76, 1.4 | 0.6 |
| ART History |  |  |  |
| Patient has never taken ARTs | — | — |  |
| Patient is taking ARTs | -0.03 | -2.8, 2.7 | >0.9 |
| Patient took ARTs but stopped | -3.7 | -8.0, 0.54 | 0.086 |

<sup>1</sup>CI = Confidence Interval

The mediation analysis found a total effect of HIV status of CT value of β = 2.3 (p < 0.001; Table 6). The indirect effect through the mediator, symptom duration, was statistically significant (β = 0.33, p = 0.048) with mediation accounting for 13.5% of the relationship. The direct effect of HIV status on Xpert CT remained statistically significant (β = 2.1, p < 0.001).

**Table 6:** Mediation Analysis of the association between HIV status and mean Xpert CT values with symptom duration as mediator among people newly diagnosed with tuberculosis in Botswana.

|  | Estimate | p-value |
| --- | --- | --- |
| Indirect Effect | 0.326 | 0.048 |
| Direct Effect | 2.085 | <0.001 |
| Total Effect | 2.411 | <0.001 |
| Prop. Mediated | 0.135 | 0.048 |

## Discussion

Using mean CT as a measure of TB bacterial burden, we found an association between decreased sputum bacterial burden and increased immunosuppression among people living with HIV in a high burden setting during the time of widespread access to ART. Additionally, increase in duration of symptoms was associated with decrease in mean CT values, indicating an increase in sputum bacterial burden. Our findings are consistent with earlier findings that those living with HIV experience lower TB sputum bacterial burden compared to those without HIV (6, 8). For example, Burger et al. compared CT values from sputum and saliva from both people living with and without HIV, and concluded that patients living with HIV had significantly lower bacterial load than individuals without HIV (17). However, prior studies did not investigate the mediating role of time to TB diagnosis. Our study findings support the observation that HIV changes the presentation of TB with a lower sputum TB bacterial load, potentially due to less inflammation and cavitation (6, 8).

As ART decreases the viral load and restores normal immune function, it has the potential to shift the TB disease presentation to be similar to those without HIV. A study using smear microscopy found that ART increased smear positivity and cavitation among participants with HIV, with lower proportions compared to individuals without HIV (18). As ART has a positive relationship with CD4 count, we expected higher CD4 counts among people with HIV in our study and similar levels of TB bacterial load by HIV status. However, our data show a significant proportion of people with HIV/TB co-infection have advanced immune suppression and, therefore, TB bacterial load remains lower among people living with HIV compared to those without HIV even in settings with widespread ART use.

Symptom duration prior to diagnosis can be used as a measure of diagnostic delay. Our data suggests a negative correlation between symptom duration and CT values after controlling for confounding variables. Previous studies suggest that people living with HIV are more likely to be diagnosed for TB faster due to more severe symptoms or increased contact with healthcare systems (9). Our data supports this finding since a significant difference was found between participants with and without HIV with symptom duration for < 1 month before diagnosis. Our findings suggest that the association between HIV-associated immunosuppression and CT values is partially mediated by differences in timeliness of TB diagnosis between people with and without HIV. However, 86.5% of this association was not mediated by TB symptom duration, which suggests that differences in pathophysiology between people with and without HIV play a dominant role in affecting TB bacterial burden.

Limitations to our study include the cross-sectional design, which limits our ability to establish causal relationships. Predictors of CT values are important to understand since the level of bacterial load has implications for transmission. Future studies should examine HIV status in relation to TB transmission among close contacts to directly demonstrate the applicability of CT values in determining transmission risks.

In conclusion, our data demonstrate that HIV status remains a predictor of TB infectiousness even when ART is widely available. Duration of symptoms was found to partially mediate this relationship. Individuals living without HIV may be at a greater risk of transmitting TB to their contacts. Further research should directly assess TB outcomes and transmission based on CT results to advance understanding of its utility in clinical and community health practice.

## Data Availability

All data produced in the present study are available upon reasonable request to the authors.

## Acknowledgements

We are grateful to the study participants who donated their time and clinical specimens to make this study possible.

